# Genomic epidemiology of Human Adenovirus F40 and F41 in Coastal Kenya: A retrospective hospital-based surveillance study (2013-2022)

**DOI:** 10.1101/2022.10.21.22281250

**Authors:** Arnold W. Lambisia, Timothy O. Makori, Martin Mutunga, Robinson Cheruiyot, Nickson Murunga, Joshua Quick, George Githinji, D. James Nokes, Charlotte J. Houldcroft, Charles N. Agoti

## Abstract

**Introduction:** Human adenoviruses type F (HAdV-F) are leading cause of childhood diarrhoeal deaths. Genomic analysis would be key for understanding their potential drivers of disease severity, transmission dynamics, and for vaccine development. However, currently there is only limited data on HAdV-F genomes globally.

**Methods:** Here, we sequenced and analysed HAdV-F from stool samples collected in coastal Kenya between 2013 and 2022. The samples were collected at Kilifi County Hospital in Kilifi, Kenya, from children < 13 years of age who reported a history of ≥ 3 loose stools in the previous 24hrs. The genomes were compared with data from the rest of the world by phylogenetic analysis and mutational profiling. Genotypes and lineages were assigned based on clustering on the global phylogenetic tree and from previously described nomenclature. Participant clinical and demographic data were linked to genotypic data.

**Results:** Of 91 cases identified using real-time PCR, 83 near-complete genomes were assembled, and these classified into HAdV-F40 and F41. These genotypes cocirculated throughout the study period. Three and four distinct lineages were observed for HAdV-F40 (Lineage 1-3) and F41 (Lineage 1, 2A, 3A, 3C and 3D). Genotype F40 and F41 coinfections were observed in five samples, and F41 and B7 in one sample. Two children with F40 and 41 coinfections were also infected with rotavirus and had moderate and severe disease, respectively. Intratypic recombination was found in 4 HAdV-F40 sequences occurring between lineages 1 and 3. None of the HAdV-F41 cases had jaundice.

**Interpretation:** This study provides evidence of extensive genetic diversity, coinfections and recombination within HAdV-F40 in a high adenovirus transmission setting that will inform public health policy, vaccine development that includes the locally circulating lineages, and molecular diagnostic assay development. We recommend future comprehensive studies elucidating on HAdV-F genetic diversity and immunity for rational vaccine development.

## Background

Human enteric adenovirus (HAdV) species F is a leading cause of paediatric viral diarrhoea and deaths globally (1). In 2019, a total of 83,491 deaths from HAdV-F were recorded globally (95% CI 43,914–143,867) in children below 14 years of age (1). Clinical isolates of HAdV-F can be classified into two genotypes (40 and 41) based on genetic differences (2,3). In Kenya, the positivity rate of HAdV-F40/41 in hospitalised <13 year-olds diarrhoeal stools was estimated at 5.8% (95% CI: 3.2-9.6) and 7.3% (95% CI: 5.2-10.1) pre- and post-rotavirus vaccine introduction, respectively (4). Cases were observed in all years studied, with peaks in dry months (5).

HAdV-F40/41 are double-stranded DNA viruses with short and long fiber proteins distinct from other adenoviruses (A-G). The genome of HAdV-F40/41 is approximately 35,000 base-pairs (6). Previously there have been HAdV studies in Ethiopia, Albania, China, India and Brazil following which the researchers have reported a higher prevalence in HAdV-F41 cases compared to F40 (7–11). Coinfections between F40 and F41, and F41 and C5 were reported in Brazil and Ethiopia (7,10). However, there are limited data on the clinical implications among individuals HAdV-F coinfections.

The emergence of new adenovirus strains due to recombination has been previously described in adenovirus species B, C and D (12–15). Intertypic recombination has not been reported in HAdV-F genotypes but intratypic recombination in HAdV F41 lineage 3b sequences has been shown to occur around the short fiber region (16). Since early 2022, HAdV-F41 has been associated with severe paediatric hepatitis, either due to triggering a dysregulated immune response that led to liver injury or through an unknown coinfection-mediated mechanism (17,18).

Understanding the genomic diversity among HAdV-F genotypes is key for optimization of molecular diagnostic assays, vaccine development, tracking global spread, and linking viral variation with disease severity. However, despite HAdV-F being among the top viral causes of diarrhoea, its genomic diversity is poorly understood due to the limited genomic data globally (∼120 as of September 2022). Notably, in Africa, there are only 10 genotype 40 and 41 complete genomes, all collected from South Africa.

Here, we aimed to investigate the molecular epidemiology and diversity of HAdV-F40/41 at the Kenyan Coast utilising 91 HAdV-F positive samples collected between January 2013 and May 2022 from children admitted to the Kilifi County Hospital (KCH) with diarrhoea.

## Methods

### Study site and population

The samples were collected as part of a prospective hospital-based rotavirus surveillance study at Kilifi County Hospital paediatric ward in Kilifi, Kenya (4). The target population was children below 13 years who presented with three or more loose stools in a 24-hour period. The samples were collected at the hospital then transported and stored at KEMRI-Wellcome Trust at -80°C.

### Ethical consideration

An informed written consent was obtained from each child’s parent/guardian before sample collection. The study protocol was approved by the Scientific and Ethics Review Unit (SERU) at Kenya Medical Research Institute, Nairobi (SERU#CGMRC/113/3624).

### Laboratory methods

#### Extraction and screening

A total of 91 real-time positive stool samples were retrieved from a -80°C storage. These included positives in samples collected between January and December 2013 and January 2016 to May 2022. The positives had been identified either by conventional real-time RT-PCR approach or by custom TaqMan Array cards as previously described (4,5). Briefly, the samples were extracted from 0.2 grams of specimen (or 200 μL if liquid) using the QIAamp Fast DNA Stool Mini kit (Qiagen, Manchester, UK) as per the manufacturer’s instructions and screened using the TaqMan Fast Virus 1-Step Master Mix and adenovirus 40/41 specific primers (Forward primer; 5’-CACTTAATGCTGACACGGGC-3’, probe; ‘FAM-TGCACCTCTTGGACTAGT-

MGBNFQ’, Reverse primer; 5’-ACTGGATAGAGCTAGCGGGC-3’). The thermocycling conditions were 95 °C for 20 sec and 35 cycles of 94 °C for 15 sec and 60 °C for 30 sec (4). No cut-off in the cycle threshold value was used during sample selection. In the HAdV-F positives, rotavirus group A was screened using conventional real-time RT-PCR approach or by custom TaqMan Array cards as previously described (4,5).

### Whole genome sequencing

#### DNA Amplification

Total nucleic acids (TNA) were amplified using the Q5® Hot Start High-Fidelity 2X Master Mix (NEB) kit. The master mix was prepared as follows: Q5® Hot Start High-Fidelity 2X Master Mix (6.25μl), H2O (3μl), Primer pool (1/2/3/4) (2μl) and TNA (1.25μl). The primers were designed by the Quick group using the ‘Jackhammer’ approach and divided the adenovirus genome into 92 amplicons of 1200bp amplified in four pools (https://github.com/quick-lab/HAdV/blob/main/HAdV-F41/v1.0/HAdV-F41_2000jh.primer.bed) (19). The reaction was then incubated on a thermocycler using the following conditions: 98°C for 30 seconds followed by 35 cycles of 98°C for 15 seconds and 65°C for 5 minutes.

### Library preparation and Sequencing

#### Oxford Nanopore Technologies (ONT)

Library preparation was performed using the SQK-LSK109 ligation kit with EXP-NBD196 barcoding kit as previously described (https://www.protocols.io/view/ncov-2019-sequencing-protocol-v3-locost-bp2l6n26rgqe/v3) (20). Briefly the amplicons were end-repaired, barcoded using EXP-NBD196 native barcoding expansion kit, pooled into one tube, adapters ligated to the library and the final library sequenced using the FLOW-MIN106D R9.4.1 flow cell on the GridION platform (Oxford Nanopore Technologies).

### Illumina Miseq

Samples that were classified as F41 based on the ONT data (described in the next section) were resequenced on the Illumina Miseq platform. An aliquot of the amplicons obtained in the DNA amplification section were used to generate libraries were generated using an Illumina library preparation kit as recommended by the manufacturer. Briefly, the amplicons were tagmented, indexed and amplified. The libraries were then normalized, pooled, and sequenced as paired end reads (2*250bp).

### Consensus genome generation and typing

Consensus HAdV-F40/41 genomes using ONT sequence data were generated using a modified primer scheme with an adenovirus reference (NC_001454) in the ARTIC field bioinformatics pipeline (https://github.com/artic-network/fieldbioinformatics) with a minimum read depth of 20x. The fast5 reads were basecalled and demultiplexed as the run progressed on the GridION machine with Guppy v.6.1.5. The reads in FASTQ format were then filtered based on the length (1200bp) and then mapped to the NC_001454 reference genome using minimap2 v.2.2.4. Variant calling and pre-consensus genome generation were performed using bcftools v.1.10.2 and then polishing and final consensus generation were performed using medaka v.1.0.3 as previously described (19).

MetaSPAdes v3.13.2 was used to de novo assemble the Illumina reads (https://github.com/ablab/spades). The reads were trimmed using QUASR v.7.03 to remove low quality reads, adapters, and primer sequences. The reads were then assembled using the default parameters on MetaSPAdes. The genomes generated from the scaffolds were then used as references and the raw reads mapped against them to check the integrity of the genomes and generate final consensus sequences.

BLASTN was used to identify the closest match for each genome in the standard nucleotide nr/nt database on NCBI (https://blast.ncbi.nlm.nih.gov/).

### Phylogenetic analysis

Only HAdV sequences with a genome coverage of ≥80% were utilized for phylogenetic analysis. There was a significant corelation between genome coverage and Ct value for HADV-F40, unlike for HAdV-F41 (**supplementary figure 1**). The sequences were compared against 111 F41 and 5 F40 genomes on GenBank as of 20^th^ September 2022 with ≥80% genome coverage (**Supplementary table 1**). Multiple sequence alignments were generated using mafft v7.487 (https://mafft.cbrc.jp/alignment/software/). The alignments were used to generate maximum likelihood (ML) phylogenies with 1000 bootstraps on IQTREE v2.1.3 (http://www.iqtree.org/) using the GTR model. The ML phylogenies were visualized using the package “ggtree” v2.4.2 (https://github.com/YuLab-SMU/ggtree) in R (https://www.r-project.org/). F40/41 genotypes were assigned based on closest hit on BLASTN and clustering of the genome on the ML tree. F41 lineages were assigned based on previously established clusters and new lineages proposed for the newly observed clusters. The F40 lineages were defined based on the clustering on the ML trees and the SNPs observed in the alignments.

### Recombination analysis

Recombination was checked using the RDP4 software and included all the detection methods i.e RDP, GENECONV, Bootscan, Maxchi, Chimaera, SiSscan, PhylPro, LARD and 3seq (21). Simplot++ (https://github.com/Stephane-S/Simplot_PlusPlus) was used to show similarity plots using a window and step size of 500bp and 100bp respectively (22). Snipit was used to visualize the mutations relative to a reference sequence (https://github.com/aineniamh/snipit).

### Statistical Analysis

All statistical analyses were performed using R version 4.1.1 (23). Comparisons among groups were made using χ^2^ statistics and a *p* value of <0.05 was considered statistically significant. Vesikari Clinical Severity Scoring System Manual was used to assess disease severity by taking into account parameters including duration and episodes of diarrhoea and vomiting, dehydration, fever, and treatment status (24). The disease severity categories were mild, moderate and severe for scores of <7, 7-10 and ≥11 respectively (24).

## Results

### Diarrhoeal cases and HAdV F genotypes in Kenya

Diarrhoea cases were documented across the years studied, with peaks observed between July and September (**Figure 1a**). Healthcare workers’ strikes interrupted sample collection in 2013, 2016 and 2017 and the COVID-19 pandemic affected sample collection in 2020 and early 2021 (25). Seventy-eight newly generated HAdV-F genomes were classified into either single genotype 40 (n=36, 43%) or 41 (n=42, 51%) samples using BLASTN and phylogenetic clustering on trees **(Figure 1b)**. Five samples had a coinfection of F40 and 41 genotypes, and one sample had a coinfection of F41 and B7 genotypes. The coinfections were investigated and confirmed by De Novo assembly and investigation of reads from both ONT and Illumina platforms. Among the five participants with the F40 and 41 coinfections, three had severe diarrhoeal disease (**Table 1**). Further, two participants with F40 and 41 coinfections were also positive for rotavirus and had moderate and severe diarrhoeal disease **(Table 1)**.

**Table 1:** Demographic characteristics of human F40/41 in children under the age of 13 years in Kilifi Kenya.

|  | F40 (n=36) | F41 (n=42) | Coinfection (n=5) | Total (n=83) | P value |
| --- | --- | --- | --- | --- | --- |
| **Gender** |  |  |  |  | 0.05 |
| Female | 21 (58.3%) | 13 (31.0%) | 2 (40.0%) | 36 (43.4%) |  |
| Male | 15 (41.7%) | 29 (69.0%) | 3 (60.0%) | 47 (56.6%) |  |
| **Age (months)** |  |  |  |  |  |
| **Median (IQR)** | 13.2 (8.2-21.6) | 15.2 (9.0-19.6) | 17.8 (16.3-21.6) | 15.2 (8.8-21.2) |  |
| **Age strata (months)** |  |  |  |  | 0.2 |
| <12 | 16 (44.4%) | 13 (31.0%) | 0 (0.0%) | 29 (34.9%) |  |
| 12-23 | 14 (38.9%) | 21 (50.0%) | 4 (80.0%) | 39 (47.0%) |  |
| 24-59 | 3 (8.3%) | 7 (16.7%) | 1 (20.0%) | 11 (13.3%) |  |
| >60 | 3 (8.3%) | 1 (2.4%) | 0 (0.0%) | 4 (4.8%) |  |
| **Disease severity** |  |  |  |  | 0.8 |
| Moderate | 17 (47.2%) | 22 (52.4%) | 2 (40.0%) | 41 (49.4%) |  |
| Severe | 19 (52.8%) | 20 (47.6%) | 3 (60.0%) | 42 (50.6%) |  |
| **Outcome** |  |  |  |  | 0.4 |
| Alive | 30 (83.3%) | 38 (90.5%) | 5 (100.0%) | 73 (88.0%) |  |
| Dead | 6 (16.7%) | 4 (9.5%) | 0 (0.0%) | 10 (12.0%) |  |
| **Rotavirus coinfection** |  |  |  |  | 0.06 |
| Positive | 2 (5.6%) | 6 (14.3%) | 2 (40.0%) | 10 (12.0%) |  |
| Negative | 34 (94.4%) | 34 (81.0%) | 3 (60.0%) | 71 (85.5%) |  |
| No data | 0 (0.0%) | 2 (4.8%) | 0 (0.0%) | 2 (2.4%) |  |

**Figure 1:**
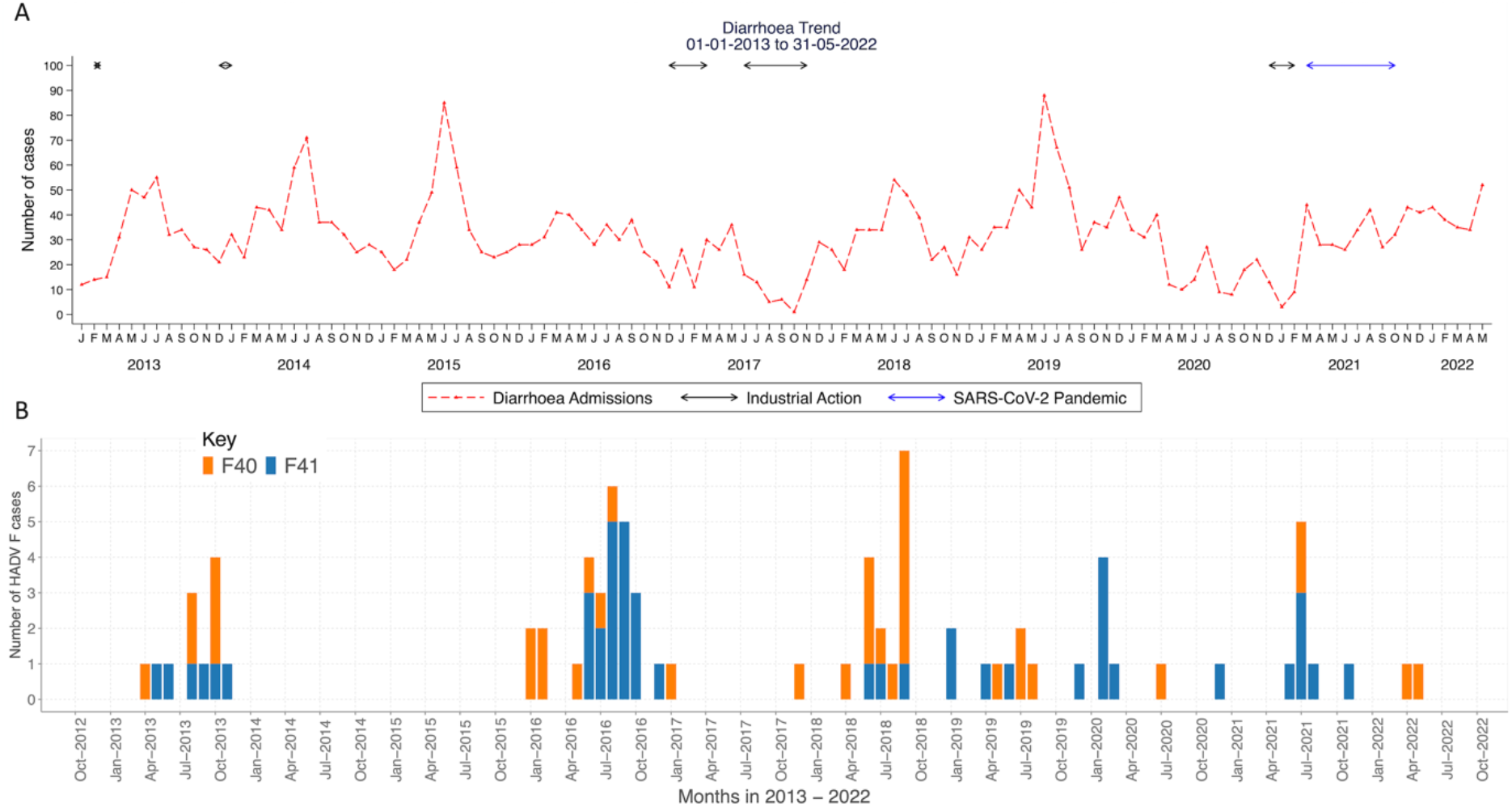
Temporal distribution of diarrhoea cases and HAdV-F40/41 cases in children under the age of 13 years in Kilifi, Kenya. **Panel a)** A line graph showing the monthly number of diarrhoea cases in Kilifi, Kenya from January 2013 to May 2022. The y-axis represents number of diarrhoea cases, and the x-axis represents months. **Panel b)** Monthly human adenovirus F40 and F41 cases observed in Kilifi, Kenya from January to December 2013, and January 2016 to May 2022. The y-axis shows the absolute number of HAdV F40 and F41 cases and the x-axis shows time in months.

There were no significant differences in gender, age strata, outcome and disease severity between the cases infected with either HAdV F40 or 41 (P > 0.05) **(Table 1)**. HAdV F40/41 and rotavirus coinfections were detected in 10 samples (12%). Of these, six were coinfections with F41 genotype (n=6). One child with HAdV-F41 and rotavirus coinfection had severe dehydration and was unconscious at the time of admission, and eventually died; the other children were treated and discharged. None of the children had jaundice as one of their illness symptoms at the time of admission.

### Diversity of HAdV-F in coastal Kenya

To determine the diversity of HAdV-F40/41 in coastal Kenya, we combined the coastal sequences with a global dataset (n=118) to give context to the newly generated genomes. We observed the clustering patterns of the coastal Kenya sequences on the global maximum likelihood phylogenetic trees and identified lineages within HAdV-F40/41. We also analysed the mutation profiles in each protein relative to a reference genome to identify synonymous and non-synonymous mutations within and between the lineages.

### HAdV-F40

Five major clusters were observed from the global phylogenetic tree (**Figure 2**). The synonymous and non-synonymous substitutions relative to the HAdV-F reference (NC_001454.1) that were characteristic for each cluster are summarized in **supplementary table 2 and supplementary figure 2**. Each of these clusters was assigned as a lineage. The Kenyan sequences, clustered with lineages 1, 2 and 3, with one global sequence observed in lineage 1 from Bangladesh (**Figure 2A**). Lineages 4 and 5 were purely non-Kenyan sequences with lineage 4 comprised of genomes from South Africa. Varied genetic divergence was observed between lineages 1 to 4 (**Supplementary figure 4**). The nucleotide percent similarity was 99·52% (nucleotide differences (nd) = 163), 99·95% (nd = 14), 99·76% (nd = 80), and 99·89% (nd = 37) for lineages 1 to 4 respectively, and 99·06% among all the five lineages.

**Figure 2:**
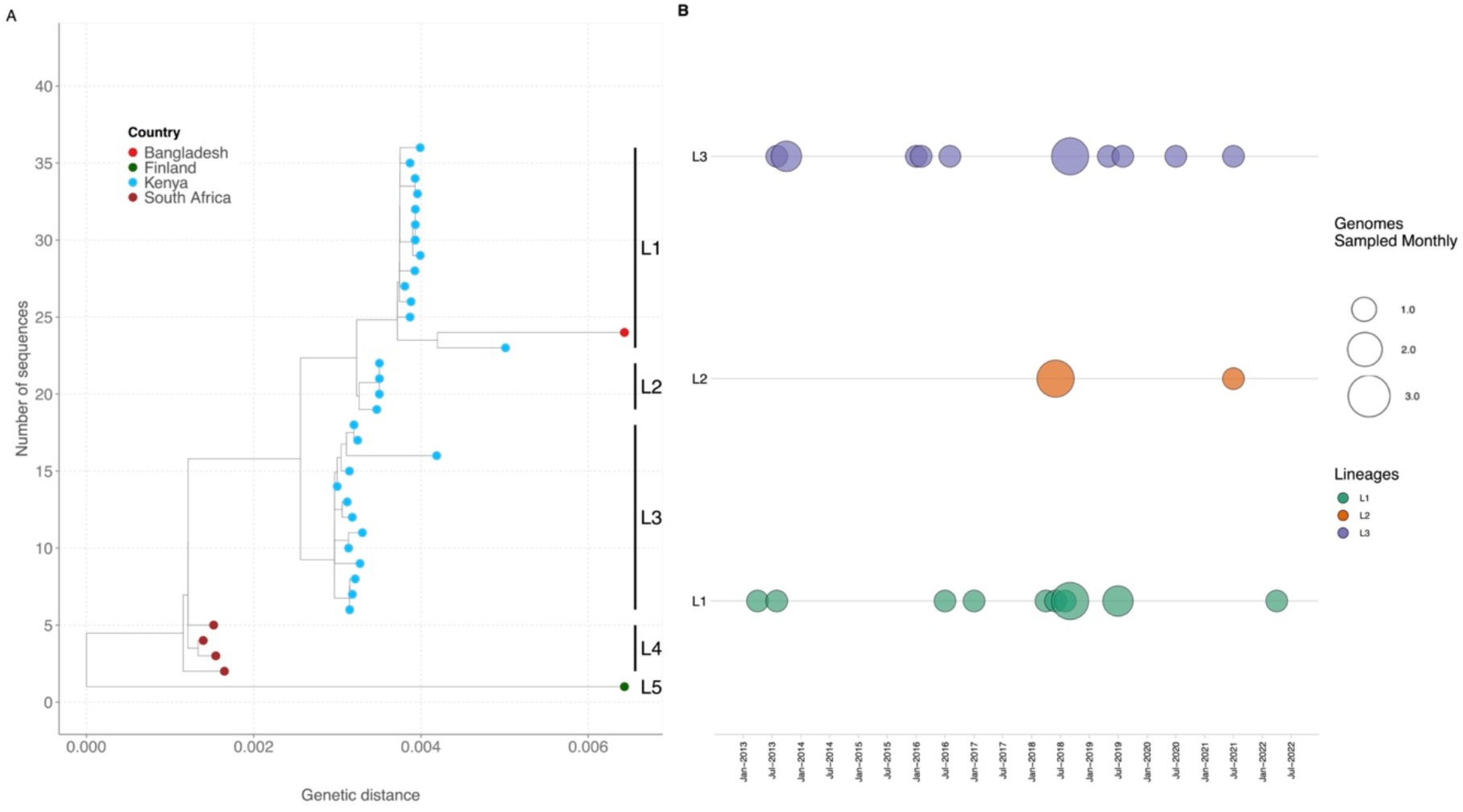
Maximum likelihood phylogenetic tree showing the genetic relatedness of HAdV-F40 genotypes when using the whole genome sequence· The tips are colored by country· **B**. Temporal distribution of HAdV-F40 lineages in sequenced samples from January to December 2013 and January 2016 to May 2022·

No amino acid differences were observed in the hexon gene of the Kenyan sequences among lineages 1 to 3 (**Supplementary table 2**). However, within the long fiber protein, lineage 2 sequences had an additional S81T amino acid mutation that was not observed lineages 1 and 3 while lineage 3 lacked the V120I mutation observed in lineage 1 and 2. Within lineage 1, sequence MN_968817.1 (Dhaka, BGD) and OP581380.1 (Kenya) contained additional showed more divergence compared to the other sequences and lacked synonymous substitutions observed within the short and long fiber proteins (**Supplementary Figure 2**). The mutation profile for the five lineages showed that lineage 2 contained polymorphic sites that contained either lineage 1 or 3 mutation profiles suggesting intertypic recombination (**Supplementary Figure 2**). Lineages 1 and 3 were in circulation throughout the study period with lineage 2 observed in 2018 and 2021 **(Figure 2B)**.

### HAdV-F41

Five lineages, 1, 2A, 2B, 3A and 3B, had been previously defined (16). The Kilifi HAdV F41 sequences clustered with global sequences characterized as lineage 1 (n=1), lineage 2A (n=14) and lineage 3A (n=4). Eleven sequences clustered within lineage 3 but neither with previously defined lineages 3A nor 3B. We propose these sequences be assigned new HAdV F41 lineages 3C and 3D (**Figure 3A**). Nonsynonymous and synonymous substitutions relative to sequence KF303070.1 (NY/2010, within lineage 2A) characteristic for each lineage were identified in multiple proteins (**Supplementary figure 3 and supplementary table 3**). A lot of the amino acid differences were observed in the hexon, 100K, E3-19·4K, E3-31·6K, E3-14·5K, short and long fiber, and E4-orf6 proteins (**Supplementary table 3**). The varied genetic divergence among the lineages has been shown in **supplementary figure 5**.

**Figure 3:**
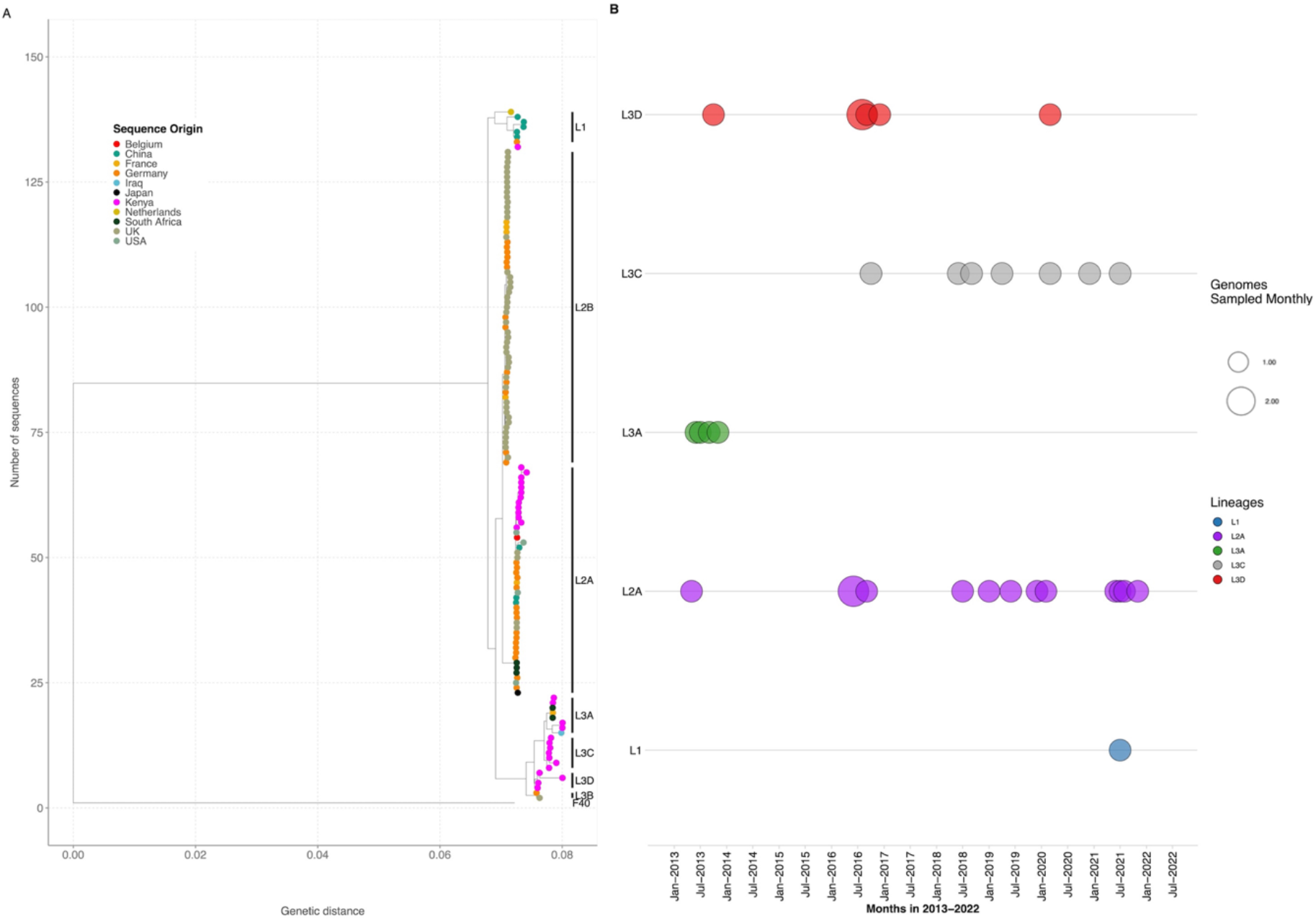
Maximum likelihood phylogenetic trees showing the diversity within the HAdV-F41 genotypes when using the whole genome sequence. The tips are colored by country. **B**. Temporal distribution of HAdV-F41 lineages in sequenced samples from January to December 2013 and January 2016 to May 2022.

In the Kenyan sequences, lineage 1 was only observed in 2021, lineage 3A in 2013, lineage 3D in 2013 and 2016, and lineage 3C from 2016 to 2021 (**Figure 3B**). Notably, lineage 2A was persistent across the study period.

The nucleotide percent identity for the newly proposed lineages is 99·87% (nd=44) and 99·85% (nd=50) for lineage 3C and 3D respectively. The percent nucleotide identity for the other lineages, 1, 2A, 2B and 3A, has been previously described and ranges from 99·6% to 99·9% (16). Within lineage diversity for each of the lineages is shown in **supplementary figure 6**.

### Recombination analysis

To investigate the possibility of recombination among the HAdV-F40 lineage 2 genomes based on the observed mutation profiles and clustering on the phylogenetic tree, we ran a recombination analysis using the RDP4 software. Two sequences were flagged as recombinants between HAdV-F40 lineage 1 (Major parent) and lineage 3 (Minor parent). The recombination was significant by RDP (p=0·00055), GENECONV (p=0.0179), Maxchi (p=0.006), Chimaera (p=0.003), SiSscan (p=0.039) and 3seq (p=0.000013). The mutation profile and similarity plot of the recombinant sequences shows switching of polymorphisms between lineage 1 and 3 sequences (**Supplementary figure 2 and 6**).

## Discussion

We report on the genomic epidemiology of HAdV F40/41 genotypes and recombination in viruses detected in hospitalised children under 13 years of age on the Kenya Coast. The observed genomic diversity has not been observed elsewhere globally. HAdV F40/41 genotypes co-circulated similar to what has been reported elsewhere (7–10). In previous studies, the proportion of F40 in sequenced HAdV-F positive samples was lower compared to compared to F41 (3-36% vs 11-97%) partly due to the higher genetic diversity in the F41 genotype that is hypothesised to enable it to outcompete F40 potentially via immune escape (7–11). A similar finding of lower F40 diversity was observed in our study. Further, the proportion of HAdV-F positive samples with a single genotype F40 infection was 41% which is higher than what has been previously described.

A coinfection between HAdV-40 and 41 has only been reported in Brazil (one sample) (10). We observed coinfections between F40 and 41 genotypes in five individuals who had moderate to severe disease (none were fatal), suggesting that coinfections do not necessarily lead to worse outcomes. Notably, reference-based genome assembly from reads in samples with coinfections generated genomes which have an anomalous phylogenetic placement, which we deconvoluted by *de novo* assembly (result not shown). Therefore, scientists and clinicians working in high adenovirus transmission settings should have a high suspicion of coinfection when they see strains which appear to be F40 × 41 recombinants and consider *de novo* assembly pipelines to pick up these coinfections.

This study contributes to the number of both HAdV-F40 and F41 genomes available on public sequence database (GenBank) four-fold. Previously, only six near complete genomes were available for HAdV-F40, our study contributes to xx additional genomes. The Kenyan HAdV-F40 sequences clustered into 3 major clusters that we defined as Lineage 1 to 3 and only one global sequence (MN968817.1-Dhaka) clustered within lineage 1. The other two lineages are purely Kenyan lineages not previously observed elsewhere (16). Over the 10-year period studied here, the co-circulation of F40 lineages 1 and 3 shows the sustained diversity of HAdV-F strains in coastal Kenya. The Kenyan HAdV-F41 genomes clustered within three previously defined lineages, 1, 2a and 3a, with none of the sequences clustering within lineage 2a and 3b, which are mainly from Europe (16). Two additional clusters composed exclusively of Kenyan sequences were observed within lineage 3 and these were not observed elsewhere globally. We have designated these two clusters as lineages 3c and 3d. Interestingly, we observe lineage replacement within the lineage 3 sub-types over time. Co-circulation and replacement of multiple HAdV-F41 strains in different genome type clusters based on the fiber gene has also been reported India between 2013 and 2020 (11,26).

HAdV-F40 lineage 2 sequences were recombinants between lineage 1 and lineage 3 sequences, and their mutation profile showed shared polymorphic sites with either of the parent lineages. The co-circulation of lineage 1 and 3 HAdV-F40 viruses could have led to the emergence of F40 recombinants. Recombination in HAdV-F40 suggests that fragment sequencing is not the best method for genotyping and lineage assignment as the recombination events may miscue visualization of the evolutionary relationships depending on the fragment sequenced.

None of the children in this study had jaundice at the time of admission between 2013 and 2022. Previous studies early in 2022 have reported human adenovirus infections among children with acute hepatitis and further studies in the coastal Kenya should investigate such cases in the future (18).

The study had two limitations. First, the genomes had amplicon dropouts meaning portions of the genome were missed and redesigning of the primers will help recover complete genomes. Finally, samples from 2014 and 2015 were not screened and tested hence some key information on the diversity of HAdV-F may have been missed from that period.

In conclusion, this study shows that there is high genetic diversity of both HAdV-F40 and F41 in co-circulation in Kenya, coupled with intratypic recombination events that may have led to the emergence of new lineages. The observed genetic diversity and recombination events necessitate the need for continuous genomic surveillance to track HAdV-F strains, to inform policy and future HAdV-F studies and develop vaccines that match what is locally circulating. In addition, the study highlights the importance of storing samples and using infrastructure from other projects, as this helps fill the knowledge gaps in the genetic diversity of pathogens through whole genome sequencing across the African continent.

## Supporting information

Supplementary figure 1, Correlation between genome coverage and the diagnostic real-time PCR cycle threshold (Ct) value for HAdV-F40 and HAdV-F41.

Supplementary figure 4. Maximum likelihood trees of HAdV-F40 lineages (A) 1, (B) 2, (C) 3 and (D) 4. The tip shapes are colored by sequence origin.

Supplementary figure 6: Percent similarity of the HAdV-F40 recombinant sequences with Lineage 1 and lineage 3 parent sequences, using SimPlot++.

Supplementary figure 5. Maximum likelihood trees of HAdV-F41 lineages (A) 1, (B) 2A, (C) 2B, (D)3A (E) 3C and (F) 3D.Tip shapes are colored by country

Supplementary figure 3, An alignment showing SNPs across the HAdV-F41 coding strand sequences relative to published sequence KF303070.1.

Supplementary figure 2, An alignment showing SNPs across the HAdV-F40 protein sequences relative to published sequence NC_001454.1.

Supplementary table 1, Information on the global reference sequences used in generating the phylogenetic tree

Supplementary table 3, Amino acid substitutions identified relative to KF303070.1 (NY/2010, within lineage 2A) in majority of sequences in F40 lineage

Supplementary table 2, Amino acid substitutions identified relative to NC_001454.1in majority of sequences in the HAdV-F40 lineages per protein.

## Data Availability

All the genomes were checked for the presence of all expected HAdV-F CDS and annotated using Geneious Prime 2022.2.1. The genomes were then deposited to GenBank with the following accession numbers: OP581345 - OP581409. The epidemiological data is available on the VEC dataverse (https://doi.org/10.7910/DVN/AMP10S).

## Contributors

Conceptualization: CNA, CJH and AWL. Methodology: AWL, JQ, CJH and CNA. Formal analysis: AWL, NM, CJH and CNA. Investigation: A.W.L, TM, RC and MM. Resources: DJN, GG, JQ and CNA. Data curation: AWL and NM. Visualization: AWL and NM. Supervision, CJH and CNA. project administration, CNA. Funding acquisition: CNA, GG and DJN. Writing—original draft preparation: AWL. All authors were involved in reviewing and editing the manuscript and agreed to submit the final version for publication.

## Data sharing

All the genomes were checked for the presence of all expected HAdV-F CDS and annotated using Geneious Prime® 2022.2.1. The genomes were then deposited to GenBank with the following accession numbers: OP581345 - OP581409. The epidemiological data is available on the VEC dataverse (https://doi.org/10.7910/DVN/AMP10S).

## Acknowledgements

We acknowledge all participants and their parents/guardians for their contribution of study samples and members of the viral epidemiology and control research group (http://virec-group.org/) for their input in the study. For the purpose of Open Access, the author has applied a CC-BY public copyright licence to any author accepted manuscript version arising from this submission.

## Funding information

This study was funded by The Wellcome Trust [102975, 203077]. Dr Charles Agoti was supported by the Initiative to Develop African Research Leaders (IDeAL) through the DELTAS Africa Initiative [DEL-407 15-003]. The DELTAS Africa Initiative is an independent funding scheme of the African Academy of Sciences (AAS)’s Alliance for Accelerating Excellence in Science in Africa (AESA) and supported by the New Partnership for Africa’s Development Planning and Coordinating Agency (NEPAD Agency). Dr. Charlotte J. Houldcroft was supported by funding from the Department of Genetics, University of Cambridge. Dr. Josh Quick was supported by funding from the UK Research and Innovation body. The views expressed in this report are those of the authors and not necessarily those of AAS, NEPAD Agency, The Wellcome, UKRI, University of Cambridge and the UK government.

## Declaration of Interest

The authors declare no competing interests.

## Supplementary data

**Supplementary table 1:** Information on the global reference sequences used in generating the phylogenetic tree.

**Supplementary table 2:** Amino acid substitutions identified relative to NC_001454.1in majority of sequences in the HAdV-F40 lineages per protein.

**Supplementary table 3:** Amino acid substitutions identified relative to KF303070.1 (NY/2010, within lineage 2A) in majority of sequences in the HAdV-F41 lineages per protein.

**Supplementary figure 1:** Correlation between genome coverage and the diagnostic real-time PCR cycle threshold (Ct) value for HAdV-F40 and HAdV-F41. The dots are colored by HAdV genotype. There was a significant corelation between genome coverage and Ct value for HAdV-F40 but not for HAdV-F41 possibly due to primer failure.

**Supplementary figure 2:** An alignment showing SNPs across the HAdV-F40 protein sequences relative to published sequence NC_001454.1. (**A**) control protein E1B 55K (**B)** capsid protein IX **(C**) E2B - DNA polymerase (**D**) Hexon protein (**E**) Short fiber **(F)** long fiber **(G)** E2B preterminal protein

**Supplementary figure 3:** An alignment showing SNPs across the HAdV-F41 coding strand sequences relative to published sequence KF303070.1 (NY/2010/4845). (**A**) Hexon protein (**B)** putative E4 ORF 6 **(C**) Long fiber (**D**) Short fiber **(E)** Penton

**Supplementary figure 4**. Maximum likelihood trees of HAdV-F40 lineages (**A**) 1, (**B**) 2, (**C)** 3 and (**D)** 4. The tip shapes are colored by sequence origin.

**Supplementary figure 5**. Maximum likelihood trees of HAdV-F41 lineages (**A**) 1, (**B**) 2A, (**C)** 2B, (**D**)3A (**E)** 3C and (**F)** 3D. The tip shapes are colored by sequence origin.

**Supplementary figure 6:** Percent similarity of the HAdV-F40 recombinant sequences with Lineage 1 and lineage 3 parent sequences, using SimPlot++ software. The color code for each HAdV-F41 parent is shown on the bottom left margin.

## Notes

### Competing Interest Statement

The authors have declared no competing interest.

### Author Declarations

The study protocol was approved by the Scientific and Ethics Review Unit (SERU) at Kenya Medical Research Institute, Nairobi(SERU#CGMRC/113/3624).

