## Supplementary figures and images for "Genomic epidemiology of Human Adenovirus F40 and F41 in Coastal Kenya: A retrospective hospital-based surveillance study (2013-2022)"

### Supplementary figure 1, Correlation between genome coverage and the diagnostic real-time PCR cycle threshold (Ct) value for HAdV-F40 and HAdV-F41.

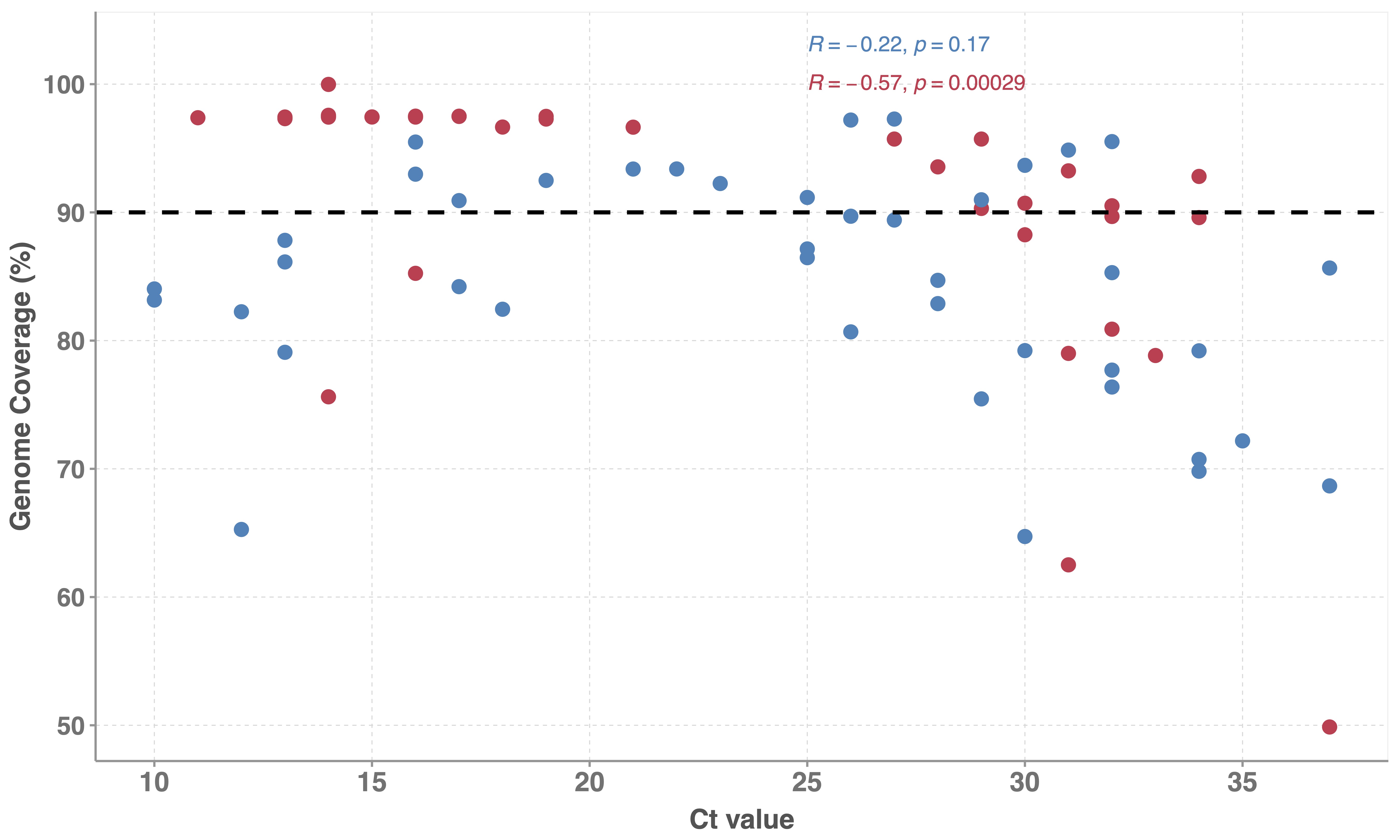

### Supplementary figure 2, An alignment showing SNPs across the HAdV-F40 protein sequences relative to published sequence NC_001454.1.

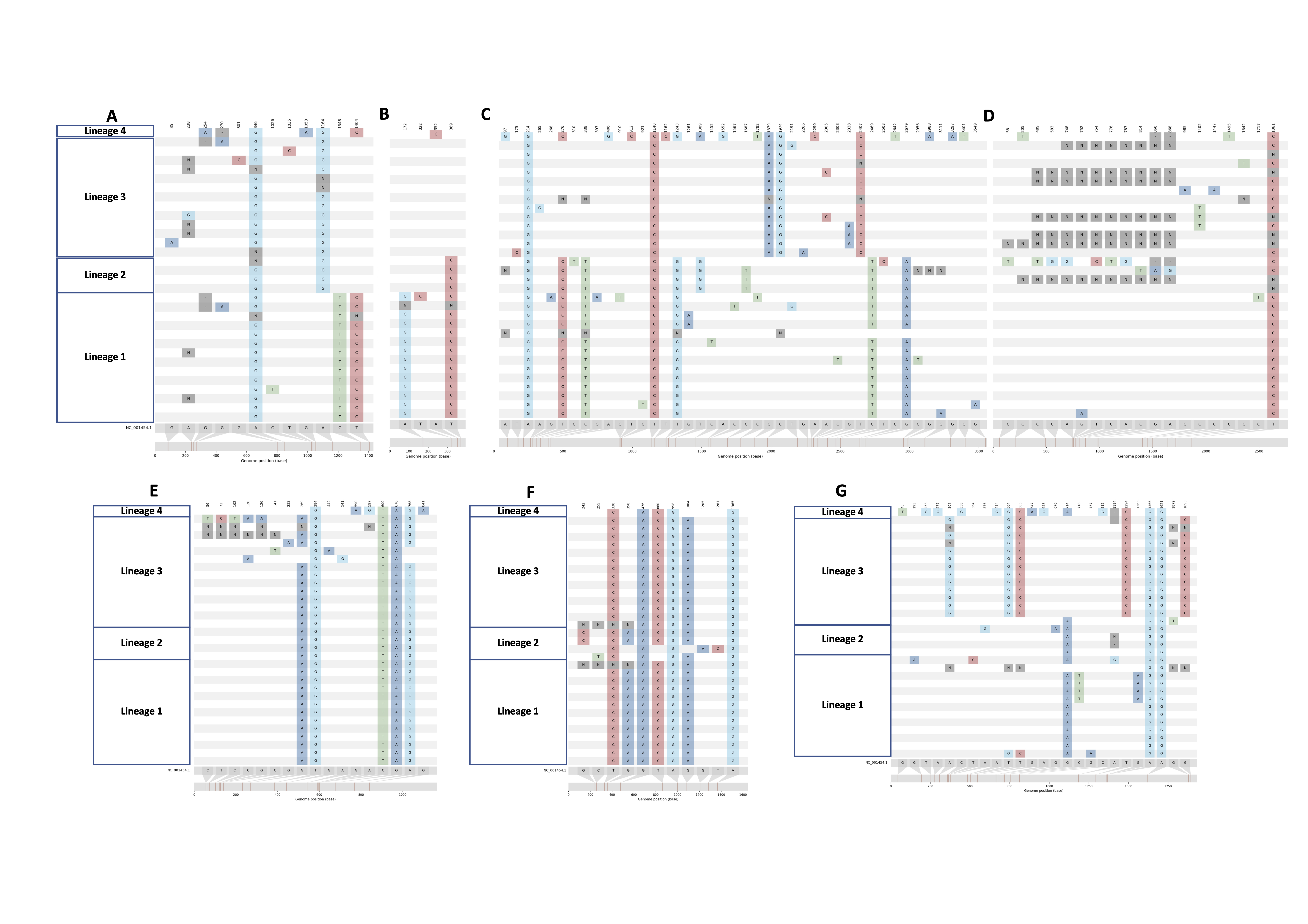

### Supplementary figure 3, An alignment showing SNPs across the HAdV-F41 coding strand sequences relative to published sequence KF303070.1.

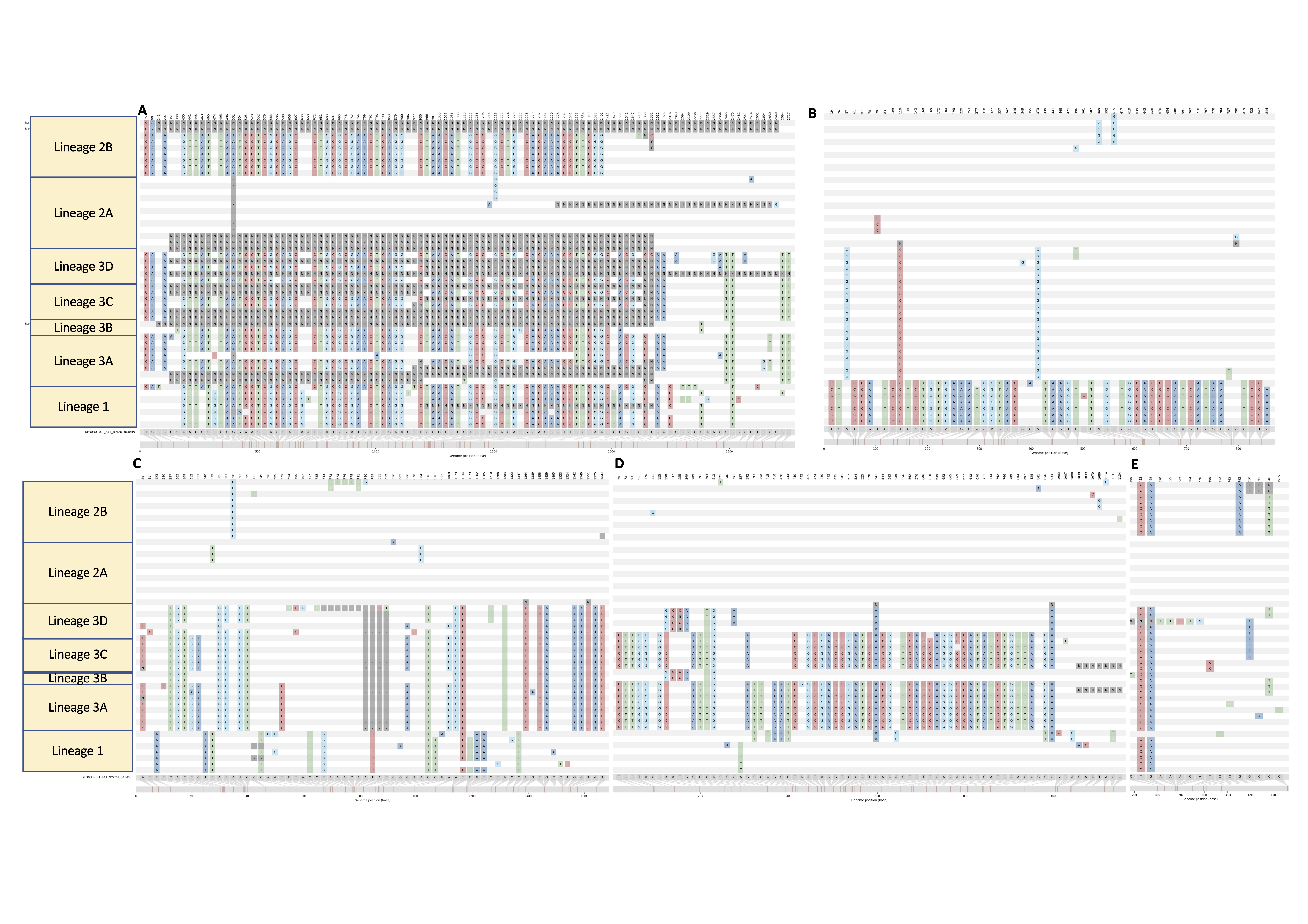

### Supplementary figure 4. Maximum likelihood trees of HAdV-F40 lineages (A) 1, (B) 2, (C) 3 and (D) 4. The tip shapes are colored by sequence origin.

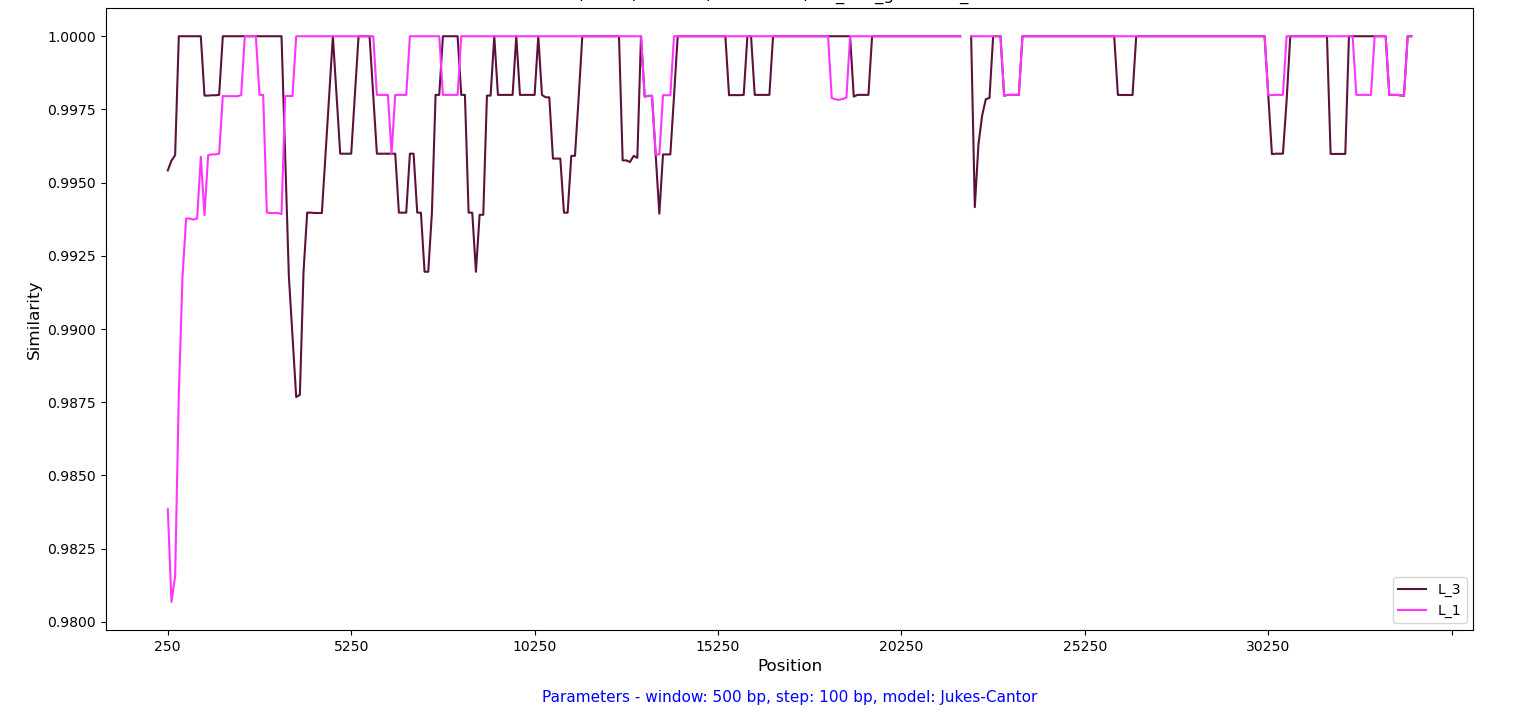

### Supplementary figure 5. Maximum likelihood trees of HAdV-F41 lineages (A) 1, (B) 2A, (C) 2B, (D)3A (E) 3C and (F) 3D.Tip shapes are colored by country

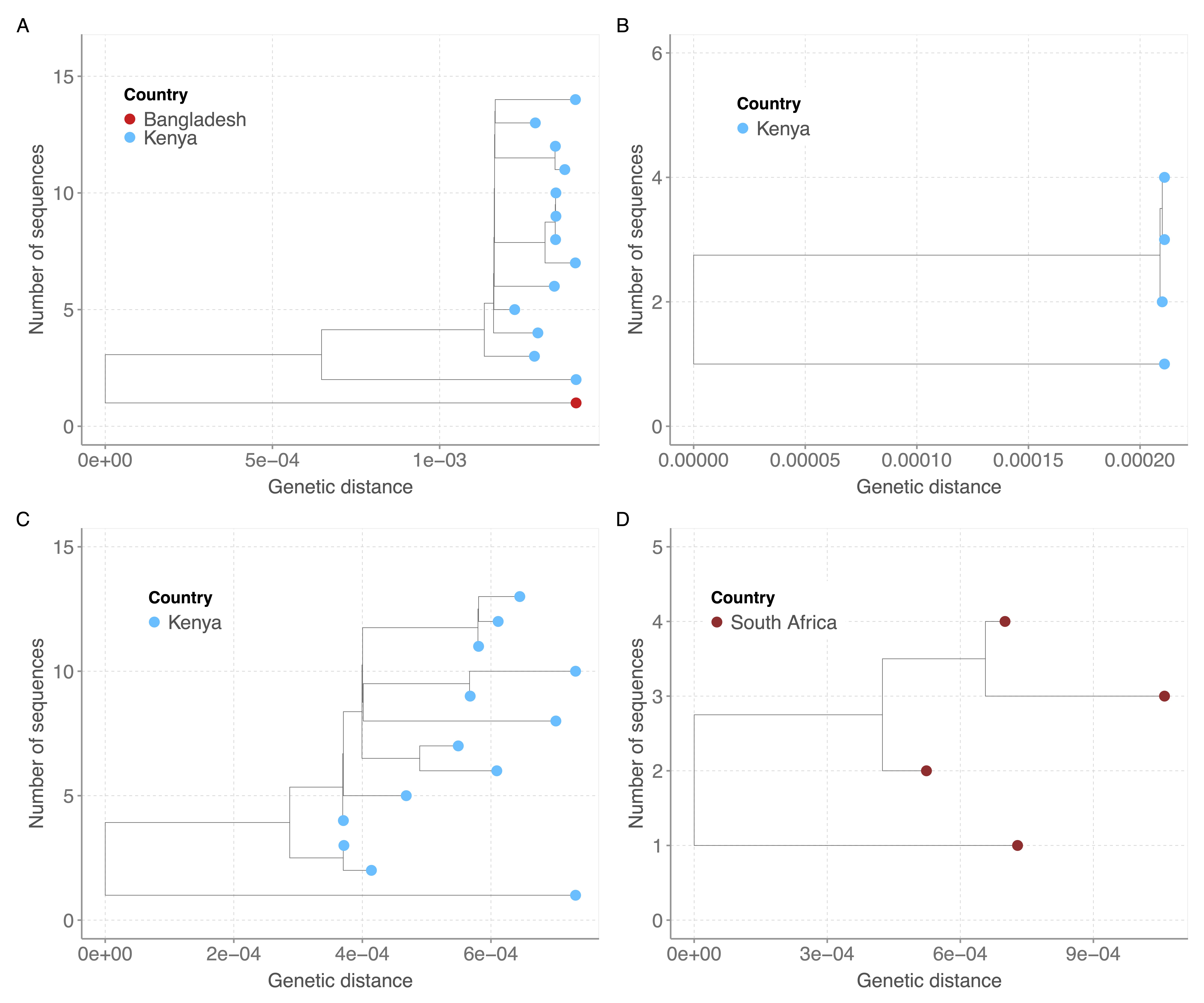

### Supplementary figure 6: Percent similarity of the HAdV-F40 recombinant sequences with Lineage 1 and lineage 3 parent sequences, using SimPlot++.

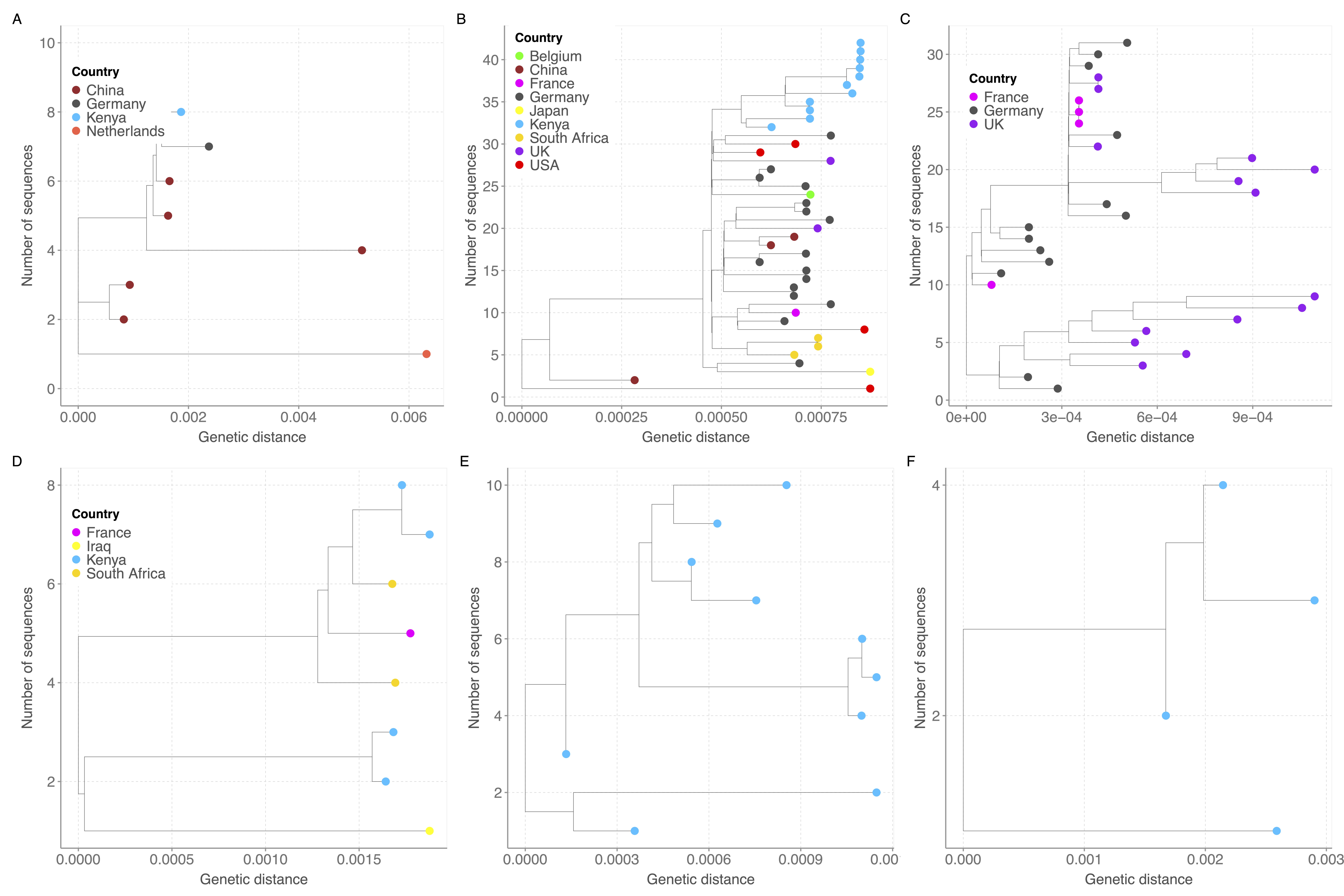
