## Supplementary table 1, Information on the global reference sequences used in generating the phylogenetic tree for "Genomic epidemiology of Human Adenovirus F40 and F41 in Coastal Kenya: A retrospective hospital-based surveillance study (2013-2022)"

| **Strain** | **Country** | **Lineage** | **Genotype** |
| --- | --- | --- | --- |
| DQ315364.2_f41Tak_Netherlands_1973 |  | L1 | F41 |
| HM565136.3_F41_NIVD103_China_2007 | China | L1 | F41 |
| KY316160.1_F41_SH/2015/D16 | China | L1 | F41 |
| KY316162.1_F41_SH/2015/D240 | China | L1 | F41 |
| KY316163.1_F41_SH/2015/D363 | China | L1 | F41 |
| MH465394.1_F41_Anhui/2018 | China | L1 | F41 |
| ON532820.1_F41_Hannover_20217_1 | Germany | L1 | F41 |
| AB728839.1_F41_SaP3-3F | Japan | L2A | F41 |
| KF303069.1_F41_NY/2010/4849 | USA | L2A | F41 |
| KF303070.1_F41_NY/2010/4845 | USA | L2A | F41 |
| KF303071.1_F41_10-4851_2010 | USA | L2A | F41 |
| KX868523.2_F41_GyK253_2000 | USA | L2A | F41 |
| KY316161.1_F41_SH/2015/D187 | China | L2A | F41 |
| KY316164.1_F41_SH/2015/D381 | China | L2A | F41 |
| MK883610.1_F41_Human/China/Shanghai/FX1-152772/2015/41 | China | L2A | F41 |
| MK962808.1_F41_SA13020_2009/2014 | South Africa | L2A | F41 |
| MK962809.1_F41_SA7335_2009/2014 | South Africa | L2A | F41 |
| MK962810.1_F41_SA12680_2009/2014 | South Africa | L2A | F41 |
| MT790999.1_F41_119-Araguaina_Brazil_2015 | Brazil | L2A | F41 |
| MT791001.1_F41_005_Porto_Nacional_BRAZIL_2015 | Brazil | L2A | F41 |
| MW567962.1_F41_MU22/patientA_France_2018 | France | L2A | F41 |
| MW686853.1_F41_Pt13_S1_2015 | UK | L2A | F41 |
| MW686857.1_F41_Pt30_S1_2019 | UK | L2A | F41 |
| MZ603083.1_HAdV-41_RVAB_2011 | Belgium | L2A | F41 |
| ON442312.1_F41_Hannover_2011_1 | Germany | L2A | F41 |
| ON442313.1_F41_Hannover_2012_1 | Germany | L2A | F41 |
| ON442314.1_F41_Hannover_2012_2 | Germany | L2A | F41 |
| ON442315.1_F41_Hannover_2012_3 | Germany | L2A | F41 |
| ON442316.1_F41_Hannover_2012_4 | Germany | L2A | F41 |
| ON442317.1_F41_Hannover_2013_1 | Germany | L2A | F41 |
| ON442318.1_F41_Hannover_2013_2 | Germany | L2A | F41 |
| ON442319.1_F41_Hannover_2013_3 | Germany | L2A | F41 |
| ON442320.1_F41_Hannover_2013_4 | Germany | L2A | F41 |
| ON442321.1_F41_Hannover_2015_1 | Germany | L2A | F41 |
| ON442322.1_F41_Hannover_2016_1 | Germany | L2A | F41 |
| ON442329.1_F41_Hannover_2022_3 | Germany | L2A | F41 |
| ON532817.1_F41_Hannover_2012_5 | Germany | L2A | F41 |
| ON532818.1_F41_Hannover_2012_6 | Germany | L2A | F41 |
| ON532819.1_F41_Hannover_2013_5 | Germany | L2A | F41 |
| ON532826.1_F41_Hannover_2022_7 | Germany | L2A | F41 |
| OP174921.1_United_Kingdom_F41_HOUA-GOSH01_2019 | UK | L2A | F41 |
| OP174922.1_United_Kingdom_F41_HOUA-GOSH02_2019 | UK | L2A | F41 |
| human/UK/Cambridge/2022/41/stool/CAM01_bc01_new/ARTIC/medaka | UK | L2B | F41 |
| human/UK/Cambridge/2022/41/stool/CAM03_bc03_new/ARTIC/medaka | UK | L2B | F41 |
| human/UK/Cambridge/2022/41/stool/CAM04_bc04_new/ARTIC/medaka | UK | L2B | F41 |
| human/UK/Cambridge/2022/41/stool/CAM05_bc05_new/ARTIC/medaka | UK | L2B | F41 |
| human/UK/Cambridge/2022/41/stool/CAM06_bc06_new/ARTIC/medaka | UK | L2B | F41 |
| human/UK/Cambridge/2022/41/stool/CAM07_bc07_new/ARTIC/medaka | UK | L2B | F41 |
| human/UK/Cambridge/2022/41/stool/CAM08_bc08_new/ARTIC/medaka | UK | L2B | F41 |
| human/UK/Cambridge/2022/41/stool/CAM09_bc09_new/ARTIC/medaka | UK | L2B | F41 |
| human/UK/Cambridge/2022/41/stool/CAM13_bc13_new/ARTIC/medaka | UK | L2B | F41 |
| human/UK/Cambridge/2022/41/stool/CAM14_bc14_new/ARTIC/medaka | UK | L2B | F41 |
| human/UK/Cambridge/2022/41/stool/CAM15_bc15_new/ARTIC/medaka | UK | L2B | F41 |
| human/UK/Cambridge/2022/41/stool/CAM21_bc21_new/ARTIC/medaka | UK | L2B | F41 |
| MG925782.1_F41_MU22_2016 | France | L2B | F41 |
| MW567964.1_F41_MU22/patientC_France_2018 | France | L2B | F41 |
| MW567965.1_F41_MU22/patientD_France_2018 | France | L2B | F41 |
| MW567966.1_F41_MU22/patientE_France_2018 | France | L2B | F41 |
| MW686854.1_F41_Pt65_S1_2019 | UK | L2B | F41 |
| MW686855.1_F41_Pt71_S1_2019 | UK | L2B | F41 |
| MW686856.1_F41_Pt63_S1_2019 | UK | L2B | F41 |
| ON442323.1_F41_Hannover_2018_1 | Germany | L2B | F41 |
| ON442324.1_F41_Hannover_2018_2 | Germany | L2B | F41 |
| ON442325.1_F41_Hannover_2021_1 | Germany | L2B | F41 |
| ON442326.1_F41_Hannover_2021_2 | Germany | L2B | F41 |
| ON442327.1_F41_Hannover_2022_1 | Germany | L2B | F41 |
| ON442328.1_F41_Hannover_2022_2 | Germany | L2B | F41 |
| ON532821.1_F41_Hannover_20217_2 | Germany | L2B | F41 |
| ON532822.1_F41_Hannover_2019_1 | Germany | L2B | F41 |
| ON532823.1_F41_Hannover_2019_2 | Germany | L2B | F41 |
| ON532824.1_F41_Hannover_2022_5 | Germany | L2B | F41 |
| ON532825.1_F41_Hannover_2022_6 | Germany | L2B | F41 |
| ON532827.1_F41_Hannover_2022_8 | Germany | L2B | F41 |
| ON561778.1_F41 | Germany | L2B | F41 |
| OP047699.1_United_Kingdom_F41_UKHSA164100247_30-Sep-16 | UK | L2B | F41 |
| OP047700.1_United_Kingdom_F41_UKHSA170480776_24-Jan-17 | UK | L2B | F41 |
| OP047701.1_United_Kingdom_F41_UKHSA180600728_30-Jan-18 | UK | L2B | F41 |
| OP047702.1_United_Kingdom_F41_UKHSA180600731_30-Jan-18 | UK | L2B | F41 |
| OP047703.1_United_Kingdom_F41_UKHSA200960804_25-Feb-20 | UK | L2B | F41 |
| OP047704.1_United_Kingdom_F41_UKHSA214680614_12-Nov-21 | UK | L2B | F41 |
| OP047705.1_United_Kingdom_F41_UKHSA214800367_23-Nov-21 | UK | L2B | F41 |
| OP047706.1_United_Kingdom_F41_UKHSA214820709_27-Nov-21 | UK | L2B | F41 |
| OP047707.1_United_Kingdom_F41_UKHSA214980732_09-Dec-21 | UK | L2B | F41 |
| OP047708.1_United_Kingdom_F41_UKHSA220200481_06-Jan-22 | UK | L2B | F41 |
| OP047709.1_United_Kingdom_F41_UKHSA220280957_13-Jan-22 | UK | L2B | F41 |
| OP047710.1_United_Kingdom_F41_UKHSA220340841_16-Jan-22 | UK | L2B | F41 |
| OP047711.1_United_Kingdom_F41_UKHSA220400725_19-Jan-22 | UK | L2B | F41 |
| OP047712.1_United_Kingdom_F41_UKHSA220400830_14-Jan-22 | UK | L2B | F41 |
| OP047713.1_United_Kingdom_F41_UKHSA220940749_23-Feb-22 | UK | L2B | F41 |
| OP047714.1_United_Kingdom_F41_UKHSA220940750_23-Feb-22 | UK | L2B | F41 |
| OP047715.1_United_Kingdom_F41_UKHSA220960496_28-Feb-22 | UK | L2B | F41 |
| OP047716.1_United_Kingdom_F41_UKHSA220960497_28-Feb-22 | UK | L2B | F41 |
| OP047717.1_United_Kingdom_F41_UKHSA221020864_06-Mar-22 | UK | L2B | F41 |
| OP047718.1_United_Kingdom_F41_UKHSA221040843_04-Mar-22 | UK | L2B | F41 |
| OP047719.1_United_Kingdom_F41_UKHSA221120661_11-Mar-22 | UK | L2B | F41 |
| OP174915.1_United_Kingdom_F41_HOUA-GOSH06_2019 | UK | L2B | F41 |
| OP174916.1_United_Kingdom_F41_HOUA-GOSH07_2019 | UK | L2B | F41 |
| OP174917.1_United_Kingdom_F41_HOUA-GOSH08_2019 | UK | L2B | F41 |
| OP174918.1_United_Kingdom_F41_HOUA-GOSH09_2019 | UK | L2B | F41 |
| OP174919.1_United_Kingdom_F41_HOUA-GOSH10_2019 | UK | L2B | F41 |
| OP174920.1_United_Kingdom_F41_HOUA-GOSH11_2019 | UK | L2B | F41 |
| OP174923.1_United_Kingdom_F41_HOUA-GOSH05_2019 | UK | L2B | F41 |
| OP174924.1_United_Kingdom_F41_HOUA-GOSH03_2019 | UK | L2B | F41 |
| OP174925.1_United_Kingdom_F41_HOUA-GOSH04_2019 | UK | L2B | F41 |
| OP174926.1_United_Kingdom_F41_HOUA-JBB27_2022 | UK | L2B | F41 |
| MG925783.1_F41_MU35_2016 | Iraq | L3A | F41 |
| MK962806.1_F41_SA13026_2009/2014 | South Africa | L3A | F41 |
| MK962807.1_F41_SA6749_2009/2014 | South Africa | L3A | F41 |
| MW567963.1_F41_MU22/patientB_France_2018 | France | L3A | F41 |
| human/UK/Cambridge/2022/41/stool/CAM02_bc02_new/ARTIC/medaka | UK | L3B | F41 |
| ON442330.1_F41_Hannover_2022_4 | Germany | L3B | F41 |
| KU162869.1_F41HoviX_1979 | Finland | L5 | F40 |
| MK955315.1_F41_SA14320_2009/2014 | South Africa | L4 | F40 |
| MK955317.1_F41_SA12730_2009/2014 | South Africa | L4 | F40 |
| MK955318.1_F41_SA12383_2009/2014 | South Africa | L4 | F40 |
| MK955319.1_F41_SA12303_2009/2014 | South Africa | L4 | F40 |
| MN968817.1_F41_human/BGD/Dhaka/2017/40 | Bangladesh | L1 | F40 |
