## Supplementary table 3, Amino acid substitutions identified relative to KF303070.1 (NY/2010, within lineage 2A) in majority of sequences in F40 lineage for "Genomic epidemiology of Human Adenovirus F40 and F41 in Coastal Kenya: A retrospective hospital-based surveillance study (2013-2022)"

**Supplementary table 3:** Amino acid substitutions identified relative to KF303070.1 (NY/2010, within lineage 2A) in majority of sequences in the HAdV-F41 lineages per protein.

| **Protein** | **L1** | **2A** | **2B** | **3A** | **3B** | **3C** | **3D** |
| --- | --- | --- | --- | --- | --- | --- | --- |
| E1A | L25P, L50Q, A111T |  |  | L25P, L50Q, A111T | L25P, L50Q | L25P, L50Q | L25P, L50Q, A111T |
| IVa2 |  |  | H37Y |  |  |  |  |
| E2B DNA polymerase | I334M, K555E |  | E305T, K555E, R709C, L725M | I334M | I334M | I334M | I334M |
| E2B preterminal protein | E288A, L362F, L598P |  |  |  |  |  |  |
| 52-55K | A332T |  |  | V125I |  | K121Q |  |
| IIIa | S465T |  |  | P456L |  | V472M, P479C |  |
| penton base |  |  | E265K |  |  |  |  |
| pVIII |  |  | A53G | A53G | A18V, A53G | A53G | A53G |
| minor core protein |  |  | I320T |  |  |  |  |
| VI |  |  | A216T |  |  | A216T |  |
| hexon | L149F, N147K, A166T, K168N, K169Q, T195A, D196Q, N228S, A250P, N251S, V255E, A266S, V285I, S409G, G410Q, T417N |  | L149F, D155N, A166T, K168N, K169Q, T195A, D196Q, N228S, A250P, N251S, V255E, A266S, V285I, S409G, G410Q, T417N | L149F, D155N, A166T, K168N, K169Q, T195A, D196Q, N228S, A250P, N251S, V255E, A266S, V285I, S409G, G410Q, T417N, T876I | L149F, D155N, A166T, K168N, K169Q, T195A, D196Q, N228S, A250P, N251S, V255E, A266S, V285I, S409G, G410Q, T417N | L149F, D155N, A166T, K168N, K169Q, T195A, D196Q, N228S, A250P, N251S, V255E, A266S, V285I, S409G, G410Q, T417N | L149F, D155N, A166T, K168N, K169Q, T195A, D196Q, N228S, A250P, N251S, V255E, A266S, V285I, S409G, G410Q, T417N |
| E2A DBP | S25P, T46A, G107D |  |  | S25P, T46A, G107D, V121I, I471V | T46A, K110N | S25P, T46A, G107D, V121I, I471V | S25P, T46A, G107D, V121I, I471V |
| 100K |  |  |  | I308V, N550S, Q570R, A695T, R737Q, C746R, A748T | I308V,N550S, Q570R, A695T, R737Q, C746R, A748T | I308V,N550S, Q570R, A695T, R737Q, C746R, A748T | I308V,N550S, Q570R, A695T, R737Q, C746R, A748T |
| pVIII | A150T |  |  | A150T | A150T | A150T | A150T |
| E3 19.4K | K17R, A22T, N33T, I144V |  |  | K17R, A22T, N33T, E44D, S48F, A75T, Q122E, L132F, Q142P, I144N, I150F, A151G, A154I, I159V, T160V, F167Y, D170E | K17R, A22T, N33T, E44D, S48F, A75T, Q122E, L132F, Q142P, I144N, I150F, A151G, A154I, I159V, T160V, F167Y, D170E | K17R, A22T, N33T, E44D, S48F, A75T, Q122E, L132F, Q142P, I144N, I150F, A151G, A154I, I159V, T160V, F167Y, D170E | K17R, A22T, N33T, E44D, S48F, A75T, Q122E, L132F, Q142P, I144N, I150F, A151G, A154I, I159V, T160V, F167Y, D170E |
| E3 31.6K | I203V, N231S, E264K |  |  | K120N, R129M, L3Q, L9F, A13V, A15V, T21N, L22F, L25P, R27Q, L36V, Q45K, D47N, S51K, N62S, N87K, T107A, H118Y, V128F, Y131H, P137S, Y146S, F181Y, Y182H, N188R, E192Q, H200Y, I203V, D218N, T22S, I245V, I246V, L253F, H257R, E264K, Q266L, Q268R | L3Q, L9F, A13V, A15V, T21N, L22F, L25P, R27Q, L36V, Q45K, D47N, S51K, N62S, N87K, T107A, H118Y, K120D*, V128F, Y131H, P137S, Y146S, F181Y, Y182H, N188R, E192Q, H200Y, I203V, D218N, T22S, I245V, I246V, L253F, H257R, E264K, Q266L, Q268R | K120N, R129M, L3Q, L9F, A13V, A15V, T21N, L22F, L25P, R27Q, L36V, Q45K, D47N, S51K, N62S, N87K, T107A, H118Y, V128F, Y131H, P137S, Y146S, F181Y, Y182H, N188R, E192Q, H200Y, I203V, D218N, T22S, I245V, I246V, L253F, H257R, E264K, Q266L, Q268R | K120N, R129M, L3Q, L9F, A13V, A15V, T21N, L22F, L25P, R27Q, L36V, Q45K, D47N, S51K, N62S, N87K, T107A, H118Y, V128F, Y131H, P137S, Y146S, F181Y, Y182H, N188R, E192Q, H200Y, I203V, D218N, T22S, I245V, I246V, L253F, H257R, E264K, Q266L, Q268R |
| E3 10.1K |  |  |  | R79T, H81R, A85T | R79T, H81R, A85T | R79T, H81R, A85T | R79T, H81R, A85T |
| E3 14.5K |  | R28H |  | I9T/P, R28H, I45L/F, L46S, I47V, G77D, L102I | I9T, R28H, I45L, L46S, I47V, G77D, L102I | I9T, R28H, I45L, L46S, I47V, G77D, L102I | I9T, R28H, I45L, L46S, I47V, G77D, L102I |
| E3 14.7K |  |  |  | K40Q | K40N | K40N |  |
| truncated U exon protein |  |  |  | D13E |  | D13E |  |
| short fiber protein |  |  |  | T62S, A97T, A128T, A140T, V165I, H173D, A175D, V181I, N182R, N190K, A217T, I218V, N212S, S226T, P228T, Y245F, H280N, L286V, L286V, D312N | S90N, | T62S, A97T, A128T, V165I, H173D, A175D, V181I, N182R, N190K, A217T, I218V, N212S, S226T, P228T, Y245F, H280N, L286V, L286V, D312N | S90N, V181I, D312N |
| long fiber protein | F250V | N295S |  | N20T, R106Q, I129V, G289D, T378A, Q427L, N255T, V487I, D517N | I129V, T378A, Q427L, N255T, V487I, D517N | N20T, R106Q, I129V, G289D, T378A, Q427L, N255T, V487I, D517N | I129V, T378A, Q427L, N255T, V487I, D517N |
| putative E4 protein 6 | Y10C, R19H, R35K, S55C, I63V, K78E, R85K, L97I, N134H, I177F, M178I, S246N, K254R, G263E, R282K |  |  | K254R, Y272H | K254R, Y272H | K254R, Y272H | K254R, Y272H |
| putative E4 protein 4 | D40N, E94K, N114S |  |  |  |  |  |  |
| putative E4 protein 3 | P70S |  |  |  |  |  |  |
