## Supplementary table 2, Amino acid substitutions identified relative to NC_001454.1in majority of sequences in the HAdV-F40 lineages per protein. for "Genomic epidemiology of Human Adenovirus F40 and F41 in Coastal Kenya: A retrospective hospital-based surveillance study (2013-2022)"

| **Protein** | **Lineage 1** | **Lineage 2** | **Lineage 3** | **Lineage 4** |
| --- | --- | --- | --- | --- |
| Control protein E1A | Y190S |  |  |  |
| Capasid protein IX | I58V |  |  | M118L |
| Encapsidation protein IVa2 | F1OL, D262N | F1OL, D262N | F1OL | F1OL |
| E2B DNA polymerase | L297F,A367T, R1077H, T1098A | L297F, A367T, R1077H, T1098A | E532Q | P56Q, L194F, R309Q, D596NE532Q,T1098A |
| E2B terminal protein precursor pTP |  |  | P17A | P632Q, G377A, H633N |
| L2 Penton |  | V24L |  |  |
| L3 Hexon | C621R | C621R | C621R | R69C, P499S, C621R |
| E3A Short fiber | S90N, G226S | S90N, G226S | S90N, G226S | G197E, G226S, D281N |
| E3A Long fiber | V120I, R159Q, M287T, N333S, A362T | S81T, V120I, R159Q, M287T, N333S, A362T | R159Q, M287T, N333S, A362T | M287T, N333S |
| E4 ORF6/7 |  |  | S51Y |  |
